# Economic burden associated with ESBL-producing *Escherichia coli* infections in Laos: econometric modeling using evidence from a prospective cost-of-illness study

**DOI:** 10.64898/2026.08.05.26358101

**Authors:** Wongyeong Choi, Phonevilai Santisouk, Yunjin Yum, Jaewoong Lee, Saehyeun Song, Palada Souvanhnavong, Khaennakhone Salodchanar, Naphaphone Khathtiyavong, Nay Thi Ha, Soulivanh Phanthavong, Loungnilanh Manivanh, Rattanaxay Phetsouvanh, Khamsay Detleuxay, Vangnakhone Dittaphong, Jung-Seok Lee

## Abstract

**Introduction:** Antimicrobial resistance poses a major global health threat, yet evidence on its economic impact in low- and middle-income countries remains limited. This study estimated the economic burden of infections caused by ESBL-producing *Escherichia coli* (*E. coli*), a key resistant pathogen, in Laos.

**Methods:** A prospective cost-of-illness study was conducted among patients with laboratory-confirmed infections in Setthathirath hospital in Vientiane, Laos, collecting cost data through repeated interviews and medical records. Descriptive analyses and econometric modeling approaches, including inverse probability weighting (IPW) and instrumental variable (IV) analyses, were used to estimate out-of-pocket expenditures, public expenditures, total cost of illness, and length of stay, accounting for potential confounding.

**Results:** ESBL-producing *E. coli* was consistently associated with higher economic burden across all analyses. The unadjusted per-patient cost was US$ 689.0 for ESBL-producing *E. coli*, compared with US$ 489.4 for non-ESBL-producing *E. coli* and US$ 537.6 for non-*E. coli*. The association remained statistically significant for out-of-pocket cost after IPW-adjustment (US$156.2; 95% CI, 14.3 to 298.1; P = 0.03), while other outcomes were not statistically significant. Instrumental variable analyses showed consistent directional effects but with wide confidence intervals and no statistically significant differences.

**Conclusions:** Findings suggest that ESBL-producing *E. coli* may be associated with increased economic burden in Laos; however, this association was not consistently statistically robust across analytical approaches. These findings suggest a potential economic impact of ESBL infection, although uncertainty remains regarding the magnitude of the effect. Strengthening antimicrobial stewardship, infection prevention and control, and improved diagnostic capacity remain essential to mitigate the potential AMR burden.

## Introduction

Antimicrobial resistance (AMR) is a major global public health concern and represents one of the most pressing challenges to modern medicine [1]. The misuse and overuse of antibiotics have made common infections increasingly difficult to treat [2]. Recent estimates identify AMR as a leading cause of death worldwide, with the greatest burden observed in low-resource settings [3]. In 2021, an estimated 4.7 million deaths were associated with AMR, including 1.1 million deaths directly attributable to resistant infections [3]. Without effective intervention, this burden is expected to increase substantially, with cumulative AMR-attributable deaths projected to reach 39.1 million globally between 2025 and 2050 [3]. Beyond its health impact, AMR imposes a substantial economic burden due to treatment failure, prolonged illness, and increased mortality [4, 5]. Projections suggest that by 2030, AMR could lead to global gross domestic product losses ranging from US$1 trillion to US$3.4 trillion [1, 6].

Among the various drivers of antibiotic resistance, infections caused by Gram-negative bacteria represent a particularly significant challenge due to their prevalence and limited treatment options [3, 8]. Gram-negative pathogens are leading causes of community- and hospital-acquired urinary tract and bloodstream infections. Among them, *Escherichia coli* (*E. coli*) is one of the most important pathogens, with resistance largely driven by extended-spectrum beta-lactamase (ESBL) production. ESBL production compromises the effectiveness of commonly used antibiotics, particularly third-generation cephalosporins [3, 8]. As a result, empirical treatment options are increasingly limited and often require the use of broad-spectrum or last-resort antibiotics, leading to a substantial burden and increased health-care costs, especially in Low- and Middle-Income countries (LMIC) [3, 8, 9].

In Laos, ESBL-producing *E. coli* was first identified in 2004, and the proportion of bacteremia caused by ESBL-producing *E. coli* has steadily increased in Vientiane [10, 11]. A retrospective study confirmed that the overall proportion of ESBL-producing *E. coli* increased nearly fivefold between 2010 to 2014 [10]. A recent study also showed that there has been a steady increase in the proportion of *E. coli* isolates that are ESBL-positive in blood cultures from 2000 to 2018, reaching 35% in 2018 [12].

Studies assessing the costs associated with ESBL-producing *E. coli* infections can generate much-needed evidence to inform infection prevention and control, antimicrobial stewardship, hospital resource allocation, and investment in diagnostics and treatment.

However, total economic impact of ESBL-producing *E. coli* infections remains insufficiently understood, particularly in LMICs, where routine cost data are often limited or unavailable [7, 13, 14]. Existing studies have also focused primarily on direct healthcare costs, with little consideration of broader components of economic burden, such as productivity loss, out-of-pocket (OOP) expenditures for non-medical sources, or costs incurred outside the primary study facility [7]. To address these gaps, this study conducted a field-based health economics study in Laos, a LMIC setting with a growing burden of ESBL-producing *E. coli* infections but limited economic data, to comprehensively evaluate the economic burden associated with ESBL-producing *E. coli* infections and generate evidence to inform policy and investment in AMR prevention and control.

## Methods

### Study design

A prospective cohort study was conducted at Setthathirath hospital, a 250-bed tertiary hospital with 16 wards located in Vientiane, Lao PDR. The hospital provides a wide range of medical and surgical services, including routine antibiotic susceptibility testing.

Data was collected from two main sources: (1) cost-of-illness (COI) surveys and (2) hospital invoices and medical records. Patients with laboratory-confirmed infections and available information on pathogen(s) and antibiotic resistance profiles were identified through hospital records and invited to participate in the study. Consented patients were enrolled and interviewed at baseline (Day 0), with follow-up interviews at Day 10–20 and at Day 30–40 if still ill. The COI surveys collected information on demographics, socioeconomic status, direct medical and non-medical costs and indirect costs. Follow-up interviews captured costs incurred since the previous interview. Interviews were conducted in person or by phone call. For child patients or those who were severely ill, interviews were conducted with a guardian or caregiver. Following completion of each interview, hospital invoices and medical records of the patient were reviewed to obtain detailed cost information and microbiological/clinical data including identified pathogen(s), antimicrobial susceptibility results, antibiotic treatment regimens, and hospitalization details.

Based on microbiological test results, enrolled patients were categorized into three cohorts: Cohort 1, patients with laboratory-confirmed detection of ESBL-producing *E. coli*; Cohort 2, patients with laboratory-confirmed detection of *E. coli* but not ESBL-producing; and Cohort 3, patients with laboratory-confirmed infections but no recorded *E. coli* bacteremia during this illness. Patients with both ESBL-producing *E. coli* and non-ESBL *E. coli* were considered as Cohort 1.

The COI analysis was conducted from societal perspective, with cost categories and valuation methods adapted from previous COI studies [15, 16]. Costs were classified into direct medical costs (DMC), direct non-medical costs (DNMC), and indirect costs (IC). DMC included consultation fees, medications, laboratory tests, and all other treatment-related expenses. To comprehensively capture DMC, hospital invoices were additionally reviewed to capture medical costs from non-patient payers. DNMC comprised non-medical expenditures incurred by patients and their companions because of the illness. These included costs for food, lodging, and transportation associated with seeking care and treatment, as well as extra payment to expedite treatment.

IC accounted for productivity losses resulting from illness, caregiver, and substitute labor. Productivity loss was estimated based on self-reported time lost from usual activities and corresponding income or school tuition (for students). For income-earning patients, productivity loss was calculated by multiplying the self-reported daily wage by the total number of workdays missed. Productivity loss was valued using the national minimum wage of Laos (2,500,000 Lao Kip per month [17]) for patients engaged in unpaid household work and self-reported school tuition for students.

In addition to patients’ own productivity losses, information was collected on the employment of substitute laborers or caregivers during the illness episode. Costs related to substitute labor or caregiving were included as reported payments (if compensated for their usual activities) or estimated opportunity costs (if unpaid and had to cut back on their usual activities).

The total COI was estimated by summing DMC, DNMC, and IC. All costs were collected in local currency (Lao Kip) and converted to United States dollars (USD) using the average annual exchange rate during 2024 [18].

### Measures

To estimate the attributable outcomes of resistance, the outcomes of Cohort 1 (ESBL-producing *E. coli*) were compared with Cohort 2 (Non-ESBL-producing *E. coli*). Furthermore, to estimate the pathogen-specific (i.e., *E. coli*) economic burden, the outcomes were compared:

Cohort 1 vs. Cohort 3 (Non-*E. coli*) who have resistant profile to at least one tested antibiotic, Cohort 2 who have resistant profile to at least one tested antibiotic vs. Cohort 3 who have resistant profile to at least one tested antibiotic, and Cohort 2 who have only susceptible profile to all tested antibiotics vs. Cohort 3 who have only susceptible profile to all tested antibiotics. For this classification, resistance was defined as non-susceptibility, with resistant or intermediate susceptibility results grouped as resistant.

### Statistical analysis

Outcomes were analyzed using descriptive statistics, as well as econometric modeling including inverse probability weighting (IPW) and instrumental variable (IV) techniques. For descriptive statistics, categorical variables were summarized by percentage and continuous variables by mean and standard deviation.

IPW was applied to minimize confounding and achieve comparability between groups with respect to measured characteristics. Propensity scores were estimated using a logistic regression model in which patient characteristics were entered to predict the probability of being in the target group (ESBL-producing *E. coli* or *E. coli*). The propensity score model included age, gender, insurance status, income group, and Charlson Comorbidity Index (CCI) score [19]. These covariates were selected based on clinical and epidemiological relevance and their potential associations with both the exposure of interest and outcomes. Patients in the target group were weighted by the inverse of the propensity score, while patients in the comparison group were weighted by the inverse of one minus the propensity score. Covariate balance before and after weighting was assessed using standardized mean differences, with an absolute value less than 0.1 indicating adequate balance.

The IV analysis is an econometric method used to mitigate potential selection bias caused by observed or unobserved confounders [20]. A valid instrument should meet two key assumptions: (1) the instrument is strongly correlated with the exposure of interest (ESBL-producing *E. coli* or *E. coli*), and (2) it affects the outcome only through the exposure, without any independent influence. In this study, the instrumental variable was defined as empirical use of selected antibiotics prior to the culture being taken. This choice was based on the assumption that initial empirical antibiotic prescribing decisions may influence the likelihood of detecting specific pathogens or resistance profiles, while not directly affecting economic outcomes except through the infection type. Specifically, for the comparison between Cohort 1 (ESBL-producing *E. coli*) and Cohort 2 (non-ESBL-producing *E. coli*), empirical use of third-generation cephalosporins prior to the *E. coli*–positive culture being taken was selected as the instrumental variable. For the comparison between Cohort 1 or 2 and Cohort 3 (non-*E. coli*), the instrumental variable was defined as empirical use of antibiotics known to be ineffective against *E. coli*, including macrolides and penicillin, administered prior to the first culture. For the IV analysis, all models were controlled for the following potential confounders: age, gender, insurance, income group, CCI score, and the presence of sepsis or urinary tract infections as disease outcomes. Two-stage residual inclusion regressions were used to evaluate the effect of the exposure of interest (ESBL-producing *E. coli* or *E. coli*) on outcomes (OOP, public expenditure, total COI burden, and length of stay (LOS)) [21-23]. In the first stage, the exposure of interest was modeled using a probit model with the instrumental variable and covariates. In the second stage, each outcome was modeled using a generalized linear model with Poisson family and log link, including the exposure of interest, covariates, and first-stage residuals, with robust standard errors. This approach accommodated non-negative and right-skewed outcomes while retaining observations with zero values in the cost outcomes [21, 23]. The relevance of the instrumental variable was assessed by examining its association with the exposure in the first-stage probit regression.

All statistical analyses including data processing, data visualization and statistical analysis were performed using Stata 18.

### Ethics statement

The COI survey questionnaires were approved by the National Ethics Committee for Health Research in Laos as well as Institutional Review Board of the International Vaccine Institute. Written informed consent or assent was obtained prior to conducting interviews, and respondents were informed that they could terminate interviews at any time. If any study participants were children under 18 years of age, assent was obtained from participants aged 12-17 years, and consent was obtained from their parents or guardians.

## Results

A total of 309 patients were enrolled in this COI study between March 28, 2025, and February 12, 2026. Of these, 73 patients had infections caused by ESBL-producing *E. coli* and were assigned to Cohort 1, while 43 patients had infections caused by non-ESBL-producing *E. coli* and were assigned to Cohort 2. The remaining 193 patients had laboratory-confirmed infections caused by non–*E. coli* pathogens and were included in Cohort 3.

Demographic characteristics by cohort were shown in Table 1. Overall, approximately 40% of patients were male among all enrolled patients, with average ages of 56.4, 55.6, and 46.9 years in Cohorts 1, 2, and 3, respectively. The mean number of sick days ranged from 16.2 to 21.2 days across cohorts with cohort 1 showing the longest illness days. Most patients had no health insurance, while about one-third were covered by public or civil insurance. Nearly all patients required a caregiver, while only small portion of patients had substitute labor. The mean self-reported household income per month was US$671.6, US$599.2, and US$746.3 in Cohort 1, 2, and 3, respectively.

**Table 1.** Descriptive statistics.

|  | Cohort 1 (SD) | Cohort 2 (SD) | Cohort 3 (SD) |
| --- | --- | --- | --- |
| Number of patients enrolled | 73 | 43 | 193 |
| % of male | 32.9% (0.5) | 30.2% (0.5) | 45.6% (0.5) |
| Average age of patients | 56.4 (20.0) | 55.6 (20.1) | 46.9 (25.2) |
| Average total number of sick days | 21.2 (9.4) | 16.2 (7.9) | 19.2 (12.8) |
| Average number of full days lost due to illness | 13.2 (8.7) | 10.0 (8.0) | 11.5 (10.0) |
| % of patients with insurance | 32.9% (0.5) | 20.9% (0.4) | 22.3% (0.4) |
| % of patients with wage loss | 27.4% (0.4) | 39.5% (0.5) | 24.9% (0.4) |
| % of patients with substitute labor | 8.2% (0.3) | 2.3% (0.2) | 2.6% (0.2) |
| % of patients with caregiver | 98.6% (0.1) | 100% (0) | 100% (0) |
| Average income per month (US\$) | 671.6 (656.2) | 599.2 (478.5) | 746.3 (798.3) |
| Disease outcomes |  |  |  |
| Pneumonia | 16.4% (0.4) | 20.9% (0.4) | 19.7% (0.4) |
| Bloodstream infections | 0% (0) | 0% (0) | 2.6% (0.2) |
| Wound or surgical site infections | 4.1% (0.2) | 2.3% (0.2) | 2.6% (0.2) |
| Sepsis | 21.9% (0.4) | 23.3% (0.4) | 18.1% (0.4) |
| Urinary tract infections | 65.8% (0.5) | 62.8% (0.5) | 18.7% (0.4) |
| Acute gastroenteritis | 2.7% (0.2) | 4.7% (0.2) | 4.7% (0.2) |
| Other diseases | 24.7% (0.4) | 20.9% (0.4) | 49.7% (0.5) |
| Charlson Comorbidity Index | 1.4 (1.5) | 0.9 (1.0) | 1.4 (1.4) |
| % of patients who were hospitalized including other facilities | 91.8% (0.3) | 100% (0) | 83.4% (0.4) |
| Average length of stay in study facility | 9.0 (6.1) | 6.3 (5.7) | 8.8 (7.7) |
| % of patients who died | 5.5% (0.2) | 7.0% (0.3) | 3.6% (0.2) |

Among disease outcomes, urinary tract infection was the most common diagnosis in both *E. coli* cohorts (Cohort 1 and 2), accounting for over 60% of cases, while a broader distribution of disease types was observed in Cohort 3. More than 90% of patients were hospitalized during the study period, and length of stay in the study facility was longest in Cohort 1 (9.0 days). During the study period, follow-up was discontinued in 14 patients due to death.

The distribution of pathogens identified from cultures obtained from COI study patients is presented in Figure 1. As expected, based on the cohort definitions, in both Cohort 1 and Cohort 2, *E. coli* was identified in all cases. In some patients, additional pathogens were identified in separate or repeated cultures from the same individuals. In contrast, a more diverse range of pathogens was observed in Cohort 3, including *Staphylococcus aureus* (36.1%) and *Klebsiella pneumoniae* (14.6%).

**Figure 1.**
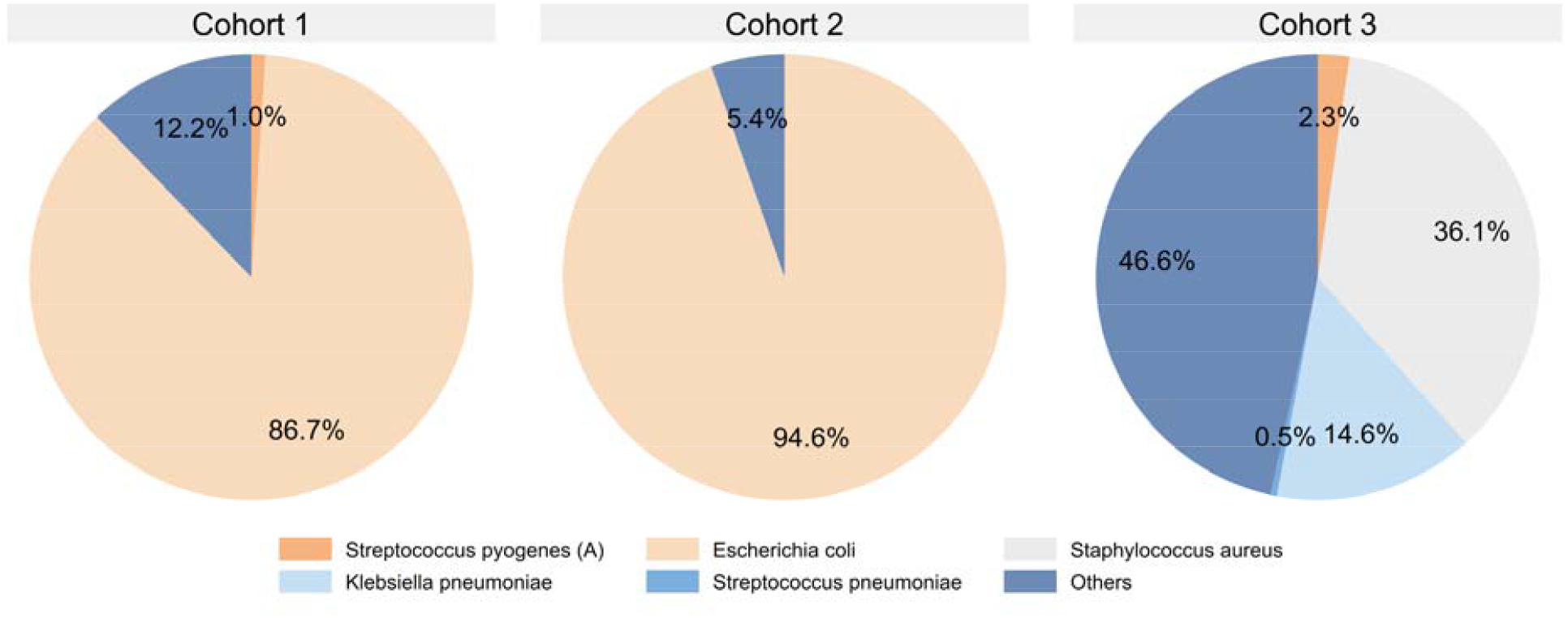
Distribution of pathogens identified from patients by cohort

Figure 2 presents the antimicrobial susceptibility test results among the study patients by cohort. Overall, the proportion of resistant cases across antibiotics was higher in Cohort 1 than in Cohort 2. Resistance to third-generation cephalosporins (Ceftriaxone and Ceftazidime) was markedly higher in Cohort 1, consistent with ESBL phenotype, whereas most isolates in Cohort 2 remained susceptible. This finding demonstrates a clear difference in resistance patterns between the two cohorts. A similar trend was observed for several other antibiotics, with Cohort 1 showing higher resistance rates compared to Cohort 2 and Cohort 3. For example, this pattern was evident for the fourth-generation cephalosporin cefepime, as well as for other antibiotic classes such as co-trimoxazole and ciprofloxacin.

**Figure 2.**
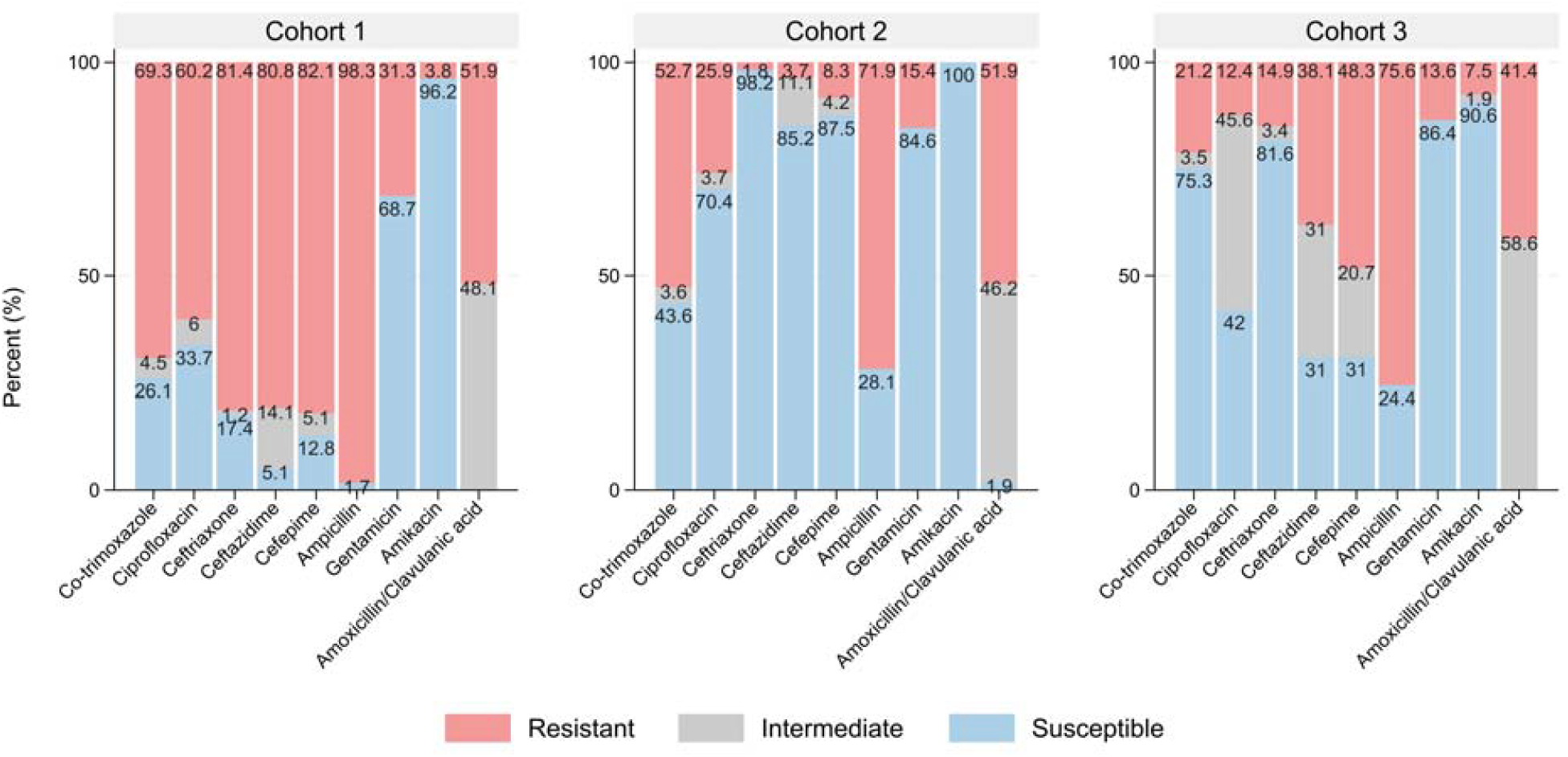
Antimicrobial susceptibility test results among the study patients by cohort

The unadjusted total mean economic burden by cohort is presented in Figure 3. As shown in Figure 3, direct medical costs accounted for approximately half of the total economic burden across all cohorts. The unadjusted total mean economic burden per patient was highest in Cohort 1 (US$ 689.0), followed by Cohort 3 (US$ 537.6) and Cohort 2 (US$ 489.4). Public expenditure accounted for approximately 6.8% to 8.3% of the total economic burden across cohorts. Indirect costs and direct non-medical costs together accounted for a substantial proportion of the remaining economic burden in all cohorts.

**Figure 3.**
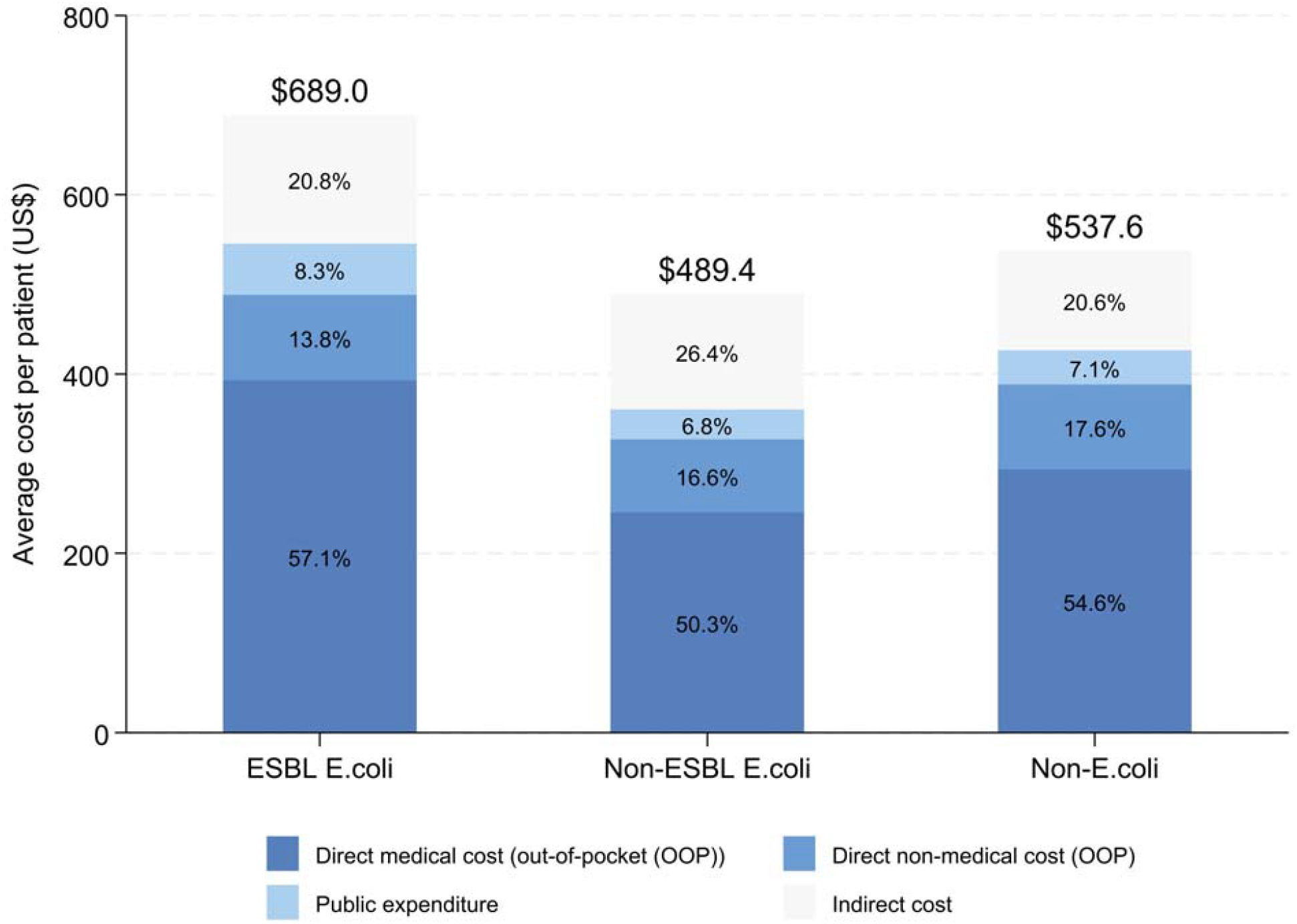
Total cost per patient by cohort and the percentage share of the economic burden by expenditure type.

The marginal effects of ESBL-producing *E. coli* and *E. coli* on economic burden using econometric modeling are presented in Figure 4 and Table 2. Across all analyses, ESBL-producing *E. coli* was consistently associated with a higher economic burden compared with non-ESBL-producing *E. coli*. In the unadjusted analysis, ESBL production was associated with an incremental out-of-pocket cost of US$161.2 and public expenditure of US$23.0. After IPW adjustment, the estimated incremental OOP costs remained statistically significant at US$156.2 (95% confidence interval [CI], 14.3 to 298.1; P = 0.03), whereas the association with public expenditure was no longer statistically significant (US$10.1, -4.5-24.8; P = 0.17). In the IV analysis, most point estimates were positive and broadly directionally consistent with the IPW findings; however, the magnitude of the IV estimates differed substantially from the IPW estimates, and most did not reach statistical significance, with wide confidence intervals throughout (Table 2). This difference may partly reflect the different estimands targeted by the two approaches, in addition to limited instrument strength in some comparisons and relatively small sample size, both of which may have further limited precision in the IV analysis (Table 2).

**Table 2.**
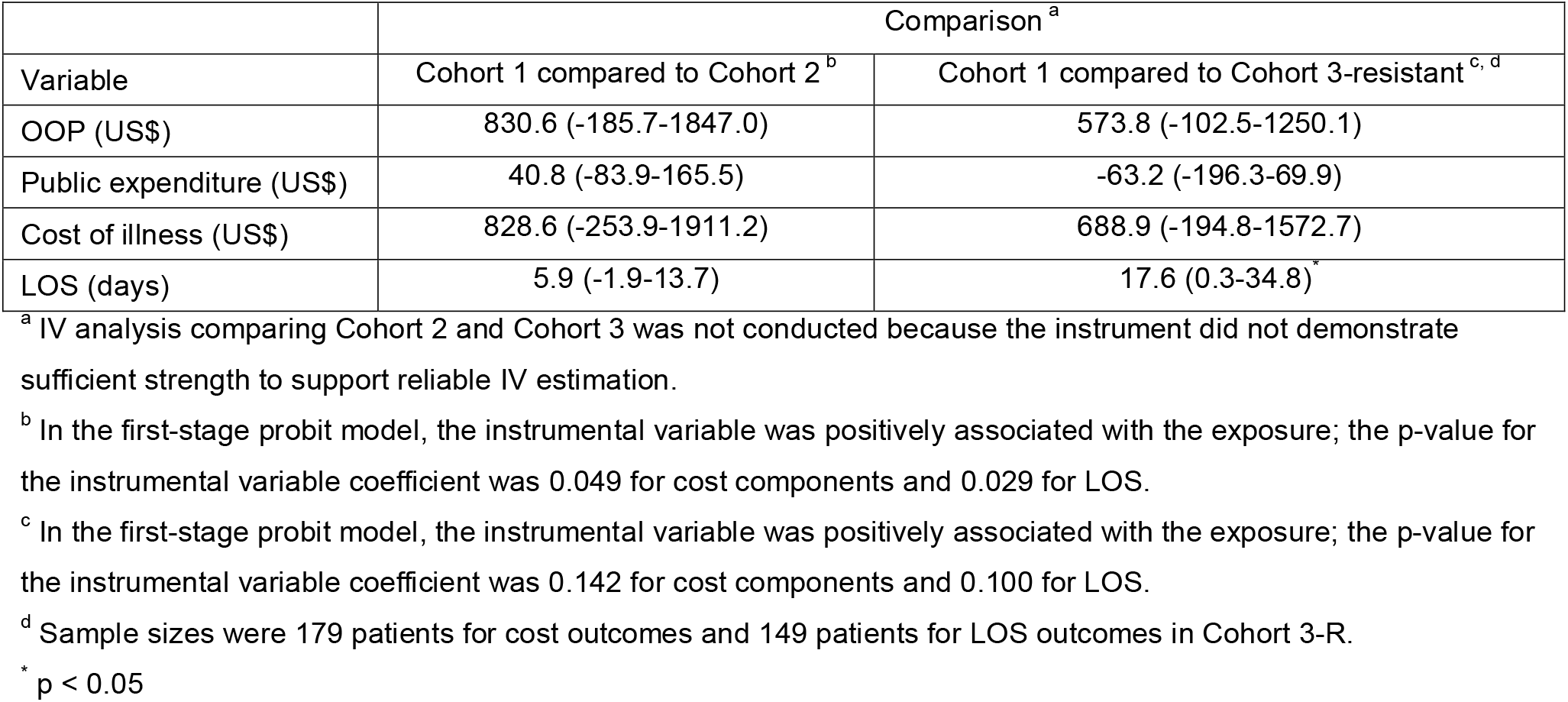
Marginal effects of ESBL-producing *E. coli* on economic burden and length of stay using the instrumental variable model.

**Figure 4.**
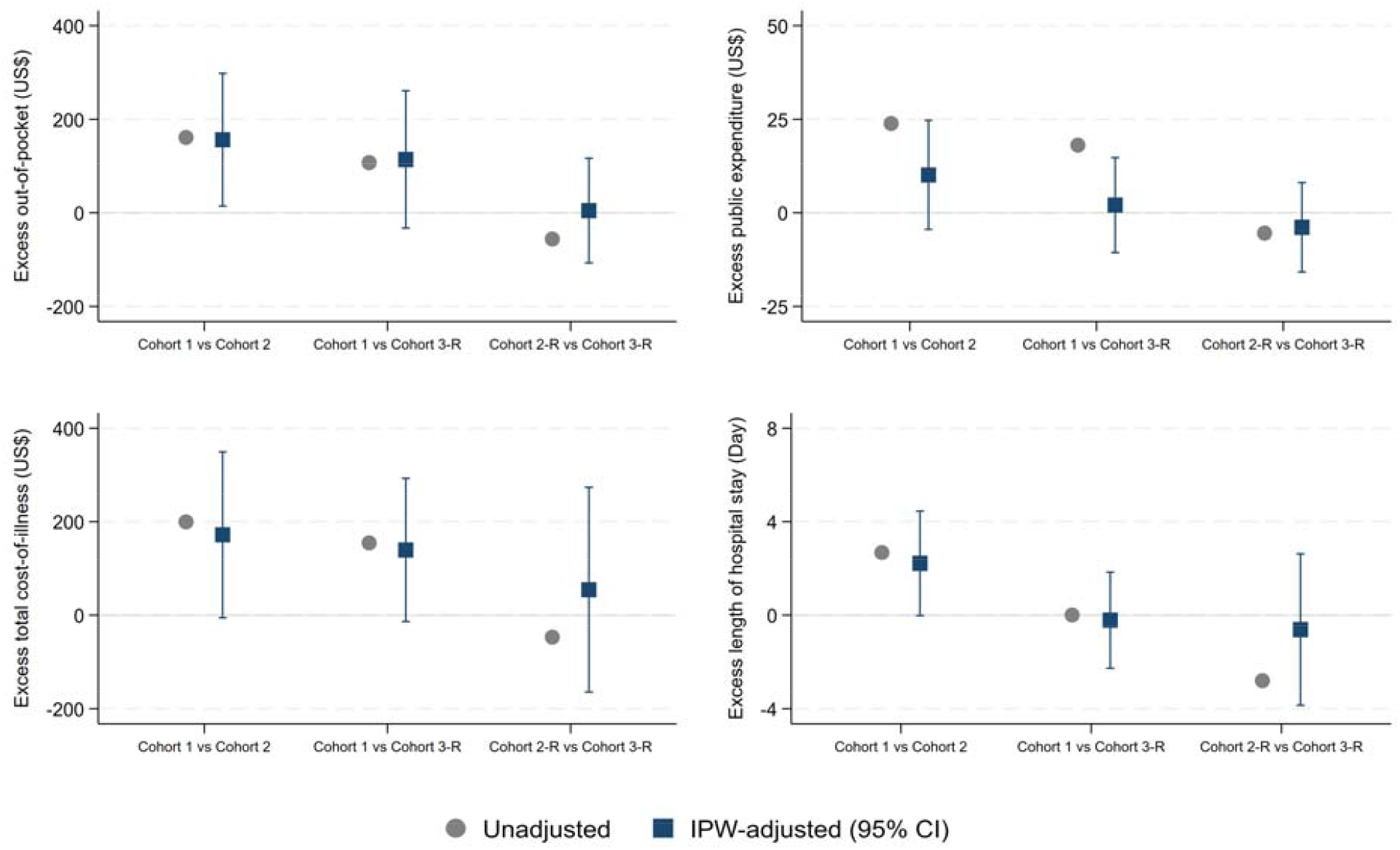
Comparison of unadjusted and IPW-adjusted excess outcomes (out-of-pocket cost, public expenditure, total cost of illness, and length of stay) Sample sizes for cost outcomes were 42 and 179 patients in Cohort 2-R and Cohort 3-R, respectively. The LOS analysis was restricted to hospitalized patients and included 67, 43, and 149 patients in Cohort 1, Cohort 2, and Cohort 3-R, respectively; among patients in Cohort 2, 42 were classified as Cohort 2-R and 1 as Cohort 2-S. An analysis comparing Cohort 2-S and Cohort 3-S was not conducted due to an insufficient sample size in Cohort 2-S.

When comparing *E. coli* cohorts to the non-*E. coli* cohort (Cohort 3), associations with economic burden were less consistent. Although unadjusted analyses suggested higher costs in Cohort 1 compared with Cohort 3, these differences were not consistently statistically significant after IPW and IV adjustments. Comparisons between Cohort 2 and Cohort 3 showed no clear evidence of increased economic burden, with estimates varying across analyses and lacking statistical significance.

## Discussion

This study conducted field-based health economics studies to quantify the impact of ESBL-producing *E. coli* by comparing the economic burden associated with those of non-ESBL-producing *E. coli* and non–*E. coli* infections. Overall, ESBL-producing *E. coli* had the highest economic burden, with an unadjusted total COI burden per patient of US$689.0, compared with US$489.4 for non-ESBL *E. coli* and US$537.6 for non-*E. coli*. Furthermore, the unadjusted LOS was longest for the ESBL-producing *E. coli* group. These findings suggest that the presence of ESBL-producing *E. coli* can be a cost driver of the increased economic burden on both patients and the healthcare system.

However, adjusted analysis yielded less consistent results after IPW-adjustment and IV analysis, suggesting that previously reported differences may be partly explained by confounding and selection bias. While the direction of the association remained consistent across analyses, substantial uncertainty remained regarding the magnitude of the estimated effect. These findings are consistent with previous studies showing mixed evidence regarding the economic impact of ESBL-producing *E. coli*. Some studies have reported that ESBL-producing *E. coli* infections are associated with increased healthcare costs and prolonged hospital stays, while others have found no significant differences in costs or clinical outcomes after adjustment for confounding factors [24-26]. Overall, these findings suggest that the economic impact of ESBL-producing *E. coli* may vary depending on study setting and analytical approach.

Compared with previous literature, this study offers several distinct advantages. First, this study captured various expenditure types including those incurred outside the healthcare system, allowing for a more comprehensive estimation of the full economic burden. Among the various cost components, DMC represented the largest private burden for patients across all cohorts, accounting for approximately half of cost of illness. Notably, DNMC (including transportation, food, and lodging) constituted a substantial portion, ranging from 13.8% to 17.6%. Similarly, IC, comprising productivity loss for patients and their caregivers or substitute labor, accounted for a substantial share, between 20.6% and 26.4%. Such expenditures are inherently difficult to capture using only data extracted from administrative healthcare databases, highlighting the importance of field-based patient surveys.

Second, this study extends beyond simple unadjusted comparisons by employing econometric modeling, enabling for a more rigorous estimation of the economic burden of AMR. Given the limited availability and quality of data in LMIC settings, the application of IV analysis and IPW was intended to enhance robustness by accounting for both observed and unobserved confounding. Although the IV analysis was constrained by limited statistical power due to the relatively small sample size, differences in marginal effects between the unadjusted and IV analyses suggest that unmeasured characteristics may have influenced the outcomes. Directional consistency across analytical approaches was observed, although statistical robustness varied.

Lastly, to the author’s knowledge, this is the first study in Laos to quantify the economic burden of AMR in Laos. While a few studies in Laos have assessed the burden of ESBL-producing *E. coli*, they have primarily focused on disease metrics such as prevalence and incidence rather than economic impact [10, 12, 27, 28]. Since most existing studies estimating the economic burden of AMR have been carried out in high-income countries, as reported by Jit et al [14] and Poudel et al [7], this study addresses an important gap by providing evidence on the economic consequences of AMR in a resource-limited setting.

Several limitations should be noted. First, the sample size was relatively small, and the study population may not fully represent the heterogeneity of the national population. In addition, as the analysis was conducted in a single hospital, caution is warranted when generalizing the findings to other settings in Laos or to similar LMIC contexts. Future studies incorporating larger and more diverse samples, as well as more detailed data, would be valuable to further validate these findings and strengthen analysis. Second, data collection relied in part on patient recall, including information from other facilities, DNMC, and IC. Third, while OOP data were captured for healthcare services used across various types of facilities, public expenditure data were limited to our facility. Therefore, public expenditure and total cost of illness may have been underestimated, although the magnitude of this underestimation is likely limited given the low coverage of health insurance in enrolled patients. Fourth, the strength of the instrumental variable varied across comparisons, with some analyses exhibiting only modest or weak associations between the instrument and the exposure. Even so, the IV model produced the estimates that were broadly consistent in direction with the other analytical approaches, supporting the overall interpretation of the results. Lastly, this study was unable to separate the economic burden attributable solely to ESBL-producing *E. coli* infections from the costs of treatment for concurrent conditions. However, by using marginal effects while holding other factors constant, the comparison between ESBL and non-ESBL infections likely captured the incremental burden associated with ESBL production.

The current study contributes to the existing knowledge on the economic burden of AMR in LMIC settings, particularly for ESBL-producing *E. coli* infections. By quantifying both public and private expenditures, this study provides evidence on the burden imposed on patients and the health system. These findings may help inform policy decisions on effective interventions such as antimicrobial stewardship programs, infection control measures, rapid diagnostic testing to support timely and appropriate antimicrobial treatment, and future vaccination strategies.

## Data Availability

Aggregated data may be made available by the corresponding author upon reasonable request and subject to the conditions of the ethical approval.

## References

1. World Health Organization. Antimicrobial resistance World Health Organization2023 [updated 21 November 2023. Available from: https://www.who.int/news-room/fact-sheets/detail/antimicrobial-resistance.

2. Iskandar K, Roques C, Hallit S, Husni-Samaha R, Dirani N, Rizk R, et al. The healthcare costs of antimicrobial resistance in Lebanon: a multi-centre prospective cohort study from the payer perspective. BMC Infect Dis. 2021;21(1):404.

3. Global burden of bacterial antimicrobial resistance 1990-2021: a systematic analysis with forecasts to 2050. Lancet. 2024;404(10459):1199–226.

4. Hillock NT, Merlin TL, Turnidge J, Karnon J. Modelling the Future Clinical and Economic Burden of Antimicrobial Resistance: The Feasibility and Value of Models to Inform Policy. Appl Health Econ Health Policy. 2022;20(4):479–86.

5. Touat M, Opatowski M, Brun-Buisson C, Cosker K, Guillemot D, Salomon J, et al. A Payer Perspective of the Hospital Inpatient Additional Care Costs of Antimicrobial Resistance in France: A Matched Case-Control Study. Appl Health Econ Health Policy. 2019;17(3):381–9.

6. Bank W. Drug-resistant infections: a threat to our economic future: World Bank; 2017.

7. Poudel AN, Zhu S, Cooper N, Little P, Tarrant C, Hickman M, et al. The economic burden of antibiotic resistance: A systematic review and meta-analysis. PLoS One. 2023;18(5):e0285170.

8. Sati H, Carrara E, Savoldi A, Hansen P, Garlasco J, Campagnaro E, et al. The WHO Bacterial Priority Pathogens List 2024: a prioritisation study to guide research, development, and public health strategies against antimicrobial resistance. Lancet Infect Dis. 2025;25(9):1033–43.

9. World Health Organization. Global antibiotic resistance surveillance report 2025. 2025 13 October 2025.

10. Chang K, Rattanavong S, Mayxay M, Keoluangkhot V, Davong V, Vongsouvath M, et al. Bacteremia Caused by Extended-Spectrum Beta-Lactamase-Producing Enterobacteriaceae in Vientiane, Lao PDR: A 5-Year Study. Am J Trop Med Hyg. 2020;102(5):1137–43.

11. Stoesser N, Crook DW, Moore CE, Phetsouvanh R, Chansamouth V, Newton PN, et al. Characteristics of CTX-M ESBL-producing Escherichia coli isolates from the Lao People’s Democratic Republic, 2004-09. J Antimicrob Chemother. 2012;67(1):240–2.

12. Roberts T, Chansamouth V, Rattanavong S, Davong V, Vongsouvath M, Mayxay M, et al. Spatio-temporal distribution of extended spectrum β-lactamase producing Escherichia coli and Klebsiella pneumoniae blood stream infections in Laos. JAC Antimicrob Resist. 2025;7(5):dlaf180.

13. Masoambeta E, Mkwanda C, Ibrahim E, Chizani K, Chapuma C, Dzanja P, et al. Economic costing methodologies for drug-resistant bacterial infections in humans in low-and middle-income countries: a systematic review. Health Econ Rev. 2025;15(1):47.

14. Jit M, Ng DHL, Luangasanatip N, Sandmann F, Atkins KE, Robotham JV, et al. Quantifying the economic cost of antibiotic resistance and the impact of related interventions: rapid methodological review, conceptual framework and recommendations for future studies. BMC Med. 2020;18(1):38.

15. Lee JS, Mogasale V, Lim JK, Carabali M, Lee KS, Sirivichayakul C, et al. A multi-country study of the economic burden of dengue fever: Vietnam, Thailand, and Colombia. PLoS Negl Trop Dis. 2017;11(10):e0006037.

16. Lee JS, Mogasale V, Lim JK, Ly S, Lee KS, Sorn S, et al. A multi-country study of the economic burden of dengue fever based on patient-specific field surveys in Burkina Faso, Kenya, and Cambodia. PLoS Negl Trop Dis. 2019;13(2):e0007164.

17. Minimum Wage – Laos WageIndicator2026 [Available from: https://wageindicator.org/salary/minimum-wage/laos.

18. World Bank. Official exchange rate (LCU per US$, period average) World Bank: World Bank; 2026 [Available from: https://data.worldbank.org/indicator/PA.NUS.FCRF.

19. Charlson ME, Pompei P, Ales KL, MacKenzie CR. A new method of classifying prognostic comorbidity in longitudinal studies: development and validation. J Chronic Dis. 1987;40(5):373–83.

20. Angrist JD, Pischke J-S. Mostly harmless econometrics: An empiricist’s companion: Princeton university press; 2009.

21. Terza JV, Basu A, Rathouz PJ. Two-stage residual inclusion estimation: addressing endogeneity in health econometric modeling. J Health Econ. 2008;27(3):531–43.

22. Biener AI, Cawley J, Meyerhoefer C. The medical care costs of obesity and severe obesity in youth: An instrumental variables approach. Health Econ. 2020;29(5):624–39.

23. Buntin MB, Zaslavsky AM. Too much ado about two-part models and transformation? Comparing methods of modeling Medicare expenditures. J Health Econ. 2004;23(3):525–42.

24. Wang Y, Xiao T, Zhu Y, Ye J, Yang K, Luo Q, et al. Economic Burden of Patients with Bloodstream Infections Caused by Extended-Spectrum β-Lactamase-Producing Escherichia coli. Infect Drug Resist. 2020;13:3583–92.

25. Gómez-Zorrilla S, RodrÍguez-Cabalé G, López Montesinos I, Alanti SS, Franquet A, Pascual-Aranda M, et al. ESBL-producing Escherichia coli bacteraemic urinary tract infections: Clinical and economic burden and antimicrobial stewardship opportunities in a retrospective cohort study. Int J Antimicrob Agents. 2026;67(6):107787.

26. Leistner R, Bloch A, Sakellariou C, Gastmeier P, Schwab F. Costs and length of stay associated with extended-spectrum β-lactamase production in cases of Escherichia coli bloodstream infection. J Glob Antimicrob Resist. 2014;2(2):107–9.

27. Chansamouth V, Mayxay M, Dance DA, Roberts T, Phetsouvanh R, Vannachone B, et al. Antimicrobial use and resistance data in human and animal sectors in the Lao PDR: evidence to inform policy. BMJ Glob Health. 2021;6(12).

28. Singh SR, Teo AKJ, Prem K, Ong RT, Ashley EA, van Doorn HR, et al. Epidemiology of Extended-Spectrum Beta-Lactamase and Carbapenemase-Producing Enterobacterales in the Greater Mekong Subregion: A Systematic-Review and Meta-Analysis of Risk Factors Associated With Extended-Spectrum Beta-Lactamase and Carbapenemase Isolation. Front Microbiol. 2021;12:695027.

